# Retinal Vascular Measures from Diabetes Retinal Screening Photographs and Risk of Incident Dementia in Type 2 Diabetes: A GoDARTS Study

**DOI:** 10.1101/2020.11.15.20230490

**Authors:** Alexander SF Doney, Aditya Nar, Yu Huang, Emanuele Trucco, Tom MacGillivray, Peter Connelly, Graham P Leese, Gareth J McKay, on behalf of the INSPIRED consortium

**Author notes:** Corresponding Author: Dr Alexander SF Doney, University of Dundee, Medical School, Division of Population Health and Genomics, Ninewells Hospital and Medical School, Dundee, Scotland, DD1 9SY, UK.

## Abstract

**Objective:** Patients with diabetes have an increased risk of dementia. Improved prediction of dementia is an important goal in developing future prevention strategies. Diabetic retinopathy screening (DRS) photographs may be a convenient source of imaging biomarkers of brain health. We therefore investigated the association of retinal vascular measures (RVMs) from DRS photographs in patients with type 2 diabetes with dementia risk.

**Research Design and Methods:** RVMs were obtained from 6,111 patients in the GoDARTS bioresource using VAMPIRE software. Their association, independent of Apo E4 genotype and clinical parameters, was determined for incident all cause dementia (ACD) and separately Alzheimer’s dementia (AD) and vascular dementia (VD). We used Cox’s proportional hazards with competing risk of death without dementia. The potential value of RVMs to increase the accuracy of risk prediction was evaluated.

**Results:** Increased retinal arteriolar fractal dimension associated with increased risk of ACD (csHR 1.17; 1.08-1.26) and AD (HR 1.33; 1.16-1.52), whereas increased venular fractal dimension (FDV) was associated with reduced risk of AD (csHR 0.85; 0.74-0.96). Conversely, FDV was associated with increased risk of VD (csHR 1.22; 1.07-1.40). Wider arteriolar calibre was associated with a reduced risk of ACD (csHR 0.9; 0.83-0.98) and wider venular calibre was associated with a reduced risk of AD (csHR 0.87; 0.78-0.97). Accounting for competing risk did not substantially alter these findings. RVMs significantly increased the accuracy of prediction.

**Conclusions:** Conventional DRS photographs could enhance stratifying patients with diabetes at increased risk of dementia facilitating the development of future prevention strategies.

## Introduction

The retina is an embryological derivative of the brain and can be imaged relatively cheaply and conveniently compared to imaging the brain. There is increasing interest in use of retinal images as a readily accessible source of imaging biomarkers for brain health (1). Such biomarkers could have clinically valuable implications, such as identifying individuals at greater risk of developing dementia for recruitment into dementia prevention trials, or as surrogate outcomes for dementia in such trials. With the failure of multiple clinical trials of potential disease-modifying treatments of dementia, possibly due to pathological changes being too advanced at patient enrolment, there is increasing interest in identifying individuals in the long prodromal phase prior to clinically manifestation of cognitive impairment for targeting prevention strategies (2). A number of studies have indicated links between retinal vascular measures (RVMs) and dementia (reviewed in 2,3). Most studies to date have been relatively small and case control in design with the possibility of confounding and reverse causation. The Rotterdam study was a large prospective study that considered pathological retinal features such as age related macular degeneration (5) and retinopathy (6) but has only considered measures of retinal vascular calibre in terms of non-pathological RVMs (7). Patients with type 2 diabetes have an increased risk of dementia compared to healthy individuals (8) and retinal screening with conventional digital photography is widely used to manage the risk of blindness due to diabetic retinopathy. Such diabetic retinopathy screening (DRS) photographs could therefore potentially also provide information about future dementia risk in patients with diabetes. Importantly, the dementias comprise a spectrum of differing and distinct pathoaetiologies dominated by the neurodegenerative features Alzheimer’s dementia (AD) and vascular dementia (VD) caused by brain injury of vascular origin, such as stroke. It is increasingly acknowledged that there is a pathological interdependence and a continuum between both major subtypes (9). Relatively few studies have compared the association of RVMs with these two major subtypes. We have recently demonstrated the ability to identify incident dementia cases using electronic medical records (EMR) in GoDARTS, a large prospective cohort linked to a genomic bioresource, and to differentiate these into VD and AD (10). In this study we obtained RVMs from the DRS photographs from patients with type 2 diabetes in GoDARTS and assessed the extent to which such measures may be used as predictors of dementia independently of other clinical and genetic risk factors.

### Study Population

GoDARTS has been previously described (11). In brief, it comprises a cohort of approximately 8500 patients who had type 2 diabetes at recruitment and 7500 individuals of similar age and background without type 2 diabetes, all of whom were recruited from the Tayside region of Scotland over a period of about 10 years commencing in 1997. Comprehensive detailed and continuously accruing EMRs are available for all participants based on an established highly deterministic regional medical information infrastructure. Since recruitment into GoDARTS began, there is now a maximum of over 20 years of follow-up data in the EMR.

## Methods

We obtained the digital DRS photographs used for the Scottish national DRS programme for all GoDARTS participants with type 2 diabetes. These photographs comprise macula-centred images with 45-degree field of view, which since the year 2000 are available as digital images. We selected the earliest available images of the right eye from each patient with acceptable quality for subjecting to measurement. Image quality was defined from DRS programme data that provides information on image gradeability. Images in which the major retinal arcade vessels were blurred, the retinal small vessels were blurred, the retinal nerve fibre layer was not visible, or where there was other technical failure were discarded. This constituted 19% of images. If the right eye image was not technically acceptable, the earliest available image from the left eye was chosen. RVMs were obtained using the Vessel Assessment and Measurement Platform for Images of the Retina (VAMPIRE) software tool (version 3; Universities of Edinburgh and Dundee, UK) which computes morphometric measurements of the retinal vasculature from a large number of retinal fundus camera images efficiently (12). The following six measures were considered for the current analysis: retinal vascular calibre as central retinal arteriole/venule equivalent (CRAE, CRVE); arteriole/venule tortuosity (TORTA/V); and arteriole/venule fractal dimension (FDV/A) which characterises the complexity of the vascular branching pattern.

We conducted a retrospective cohort study of the association of this set of RVMs with incident dementia in GoDARTS EMR using our previously described and validated methodology (10). Cox’s proportional hazards was used with entry time being the date of acquisition of the DRS photograph that was measured by VAMPIRE and exit time being the date of first available evidence in the EMR for a diagnosis of dementia, which, as previously, was considered as a surrogate for dementia incidence. Censoring was end of available follow-up EMR data or death without EMR evidence of dementia. In addition to ACD we considered AD and VD separately as defined by our previous analysis. We did not separately evaluate the relatively small numbers of individuals who either could not be adjudicated as having AD or VD or who had a rarer cause of dementia. Apo E4 status defined by rs429358 genotype was included in all analyses and modelled as a codominant co-variable. The following clinical co-variables were also included in all models: sex (females coded as 1, males 0), age, smoking status (as ever/never smoked), years with diabetes, a history of hospitalisation with major adverse cardiovascular event (previous CVD), body mass index (BMI), systolic blood pressure (SBP), total cholesterol, high density lipoprotein cholesterol (HDL) and glycated haemoglobin. Continuous clinical covariables were evaluated as mean of all available values during a 3-year period prior to date of study entry. Where these values were missing for a particular individual the value was imputed to the overall cohort mean value. Due to their potential for interdependence, all RVMs and all clinical covariates were included in all models with stepwise backward selection with p=0.2 as the selection threshold for staying in the final model. To aid meaningful comparisons, all RVMs were z-standardized so hazard ratios (HR) associated with RVMs are all provided per standard deviation (SD) increase.

As age is the major determinant of dementia risk, and RVMs have been previously associated with features of cardiovascular diseases and aging (13), we set the time scale for the analysis as years of age, meaning all models were continuously adjusted for age. We also incorporated a competing risk of death without dementia analysis where risk is expressed as a sub-distribution HR (sdHR). All HR are expressed with their 95% confidence intervals. As this was an incident study, any individuals who had an established dementia diagnosis in their EMR on or prior to the date of study entry were excluded. Finally, to evaluate the potential of RVMs as biomarkers for dementia, we performed receiver operator characteristic (ROC) analyses. In this case, for practicality of comparing area under the curve (AUC) between models, we undertook logistic regression to predict risk of dementia within 10 years. We compared AUC for predicting ACD, VD and AD using clinical variables and ApoE4 genotype alone with AUCs in which RVMs were also added. For further assessment of the potential clinical value of RVMs we determined the Net Reclassification Improvement (NRI) and Integrated Discrimination Improvement (IDI) (14). For NRI we set the risk threshold at 10% and 20% with the motivation being that the 10-year risk of dementia in this population was on average approximately 10%. As has been previously recommended (15), to improve interpretation of the NRI we also determined its sub-components in terms of the NRI for events (NRI*e*) and non-events (NRI*ne*) separately. STATA 14 was used for all data assembly and analyses. The *nriidi* package developed by Sundström *et al* (16) was used to evaluate IDI and NRI.

## Results

A total of 6111 patients with type 2 diabetes had RVMs successfully measured and had a full set of data for analysis. The baseline characteristics of this population are provided in Table 1. Figure S1 provides a correlation heatmap of all the covariates used in this study. The mean age at study entry was 68.4 years (IQR 60-75.5) with a mean duration of diabetes of 7.1 years (IQR 7.1-11.8).

**Table 1:** Study Population Baseline Characteristics.

|  |  |
| --- | --- |
| **Total population** | 6111 |
| **Males %** | 56% |
| **Mean Age at measured DRS photograph (IQR) years** | 68.4 (60.1-75.5) |
| **Mean duration diabetes (IQR) years** | 7.1(3.8-11.8) |
| **Previous CVD event** | 28.3% |
| **Mean Glycated Hb (SD) %/mmolmol<sup>-1</sup>** | 7.52% (1.11)/ 59.0 (12.1) |
| **Mean Total Cholesterol (SD) mmolL<sup>-1</sup>** | 4.37(0.84) |
| **Mean HDL Cholesterol mmolL<sup>-1</sup>** | 1.34(0.35) |
| **Mean BMI kgm<sup>-2</sup>** | 31.13(5.97) |
| **Mean SBP mmHg** | 139(11.5) |
| **Smoking** | 50% |
| **Apo e4 (rs429358) genotypes (%)** | TT - 72.2%<br>TC - 24.9%<br>CC- 2.9% |
| **CRAE (SD) pixels** | 32.6 (2.7) |
| **CRVE (SD) pixels** | 42.9 (3.5) |
| **Log TORTA (SD)** | -9.79 (0.98) |
| **Log TORTV (SD)** | -9.71 (0.77) |
| **FDA(SD)** | 1.51 (0.07) |
| **FDV(SD)** | 1.51 (0.07) |

Fifty-nine individuals were excluded as already having a record for dementia in the EMR at the date of acquisition of the retinal photograph. The median follow-up time in the remaining 6052 individuals was 9.8 years (IQR 5.7-10.6) during which there were 635 incident cases of ACD comprising 327 (51.5%) AD, 218 (34.3%) VD and 90 (14.2%) undeterminable dementia type or other rarer dementia diagnoses. The overall incidence rate was 13 (lower bound 12 upper bound 14) per 1000 person years. There were 1844 deaths without evidence dementia in the EMR at or prior to date of death.

Table 2 provides the cause specific-hazard ratios (csHR) for the Cox’s regression model and the sub-distribution (sdHR) for the competing risk model for the final set of variables remaining after backward stepwise removal for each of ACD, AD and VD. For ACD we found a significant independent association of increased arteriolar fractal dimension (FDA) with increased risk of incident ACD (csHR 1.17; 1.08-1.26). Accounting for competing risk of death resulted in little change (sdHR 1.16; 1.08-1.25). Furthermore, wider arteriolar calibre (CRAE) was associated with a reduced risk (csHR 0.9; 0.83-0.98) although this was no longer significant when considering competing risk of death (sdHR 0.93; 0.86-1.00).

**Table 2:** Covariates remaining in the Cox and Competing risk models after stepwise removal for ACD, AD and VD (-denotes removed covariable)

| ACD | Cox Model |  |  | Competing Risk model |  |  |
| --- | --- | --- | --- | --- | --- | --- |
|  | csHR | CI | p | sdHR | CI | p |
| Age | 0.93 | 0.91-0.96 | <0.000 | 1.11 | 1.09-1.22 | <0.000 |
| Sex | - | - | - | 1.19 | 1.02-1.39 | 0.032 |
| Duration with T2D | 1.02 | 1.01-1.03 | <0.000 | 1.01 | 1.00-1.02 | 0.091 |
| Previous CVD | - | - | - | 0.82 | 0.69-0.98 | 0.025 |
| GHB | 1.13 | 1.04-1.23 | 0.004 | - | - | - |
| SBP | 0.99 | 0.99-1.00 | 0.08 | 0.99 | 0.99-1.00 | 0.164 |
| TC | - | - | - | 1.09 | 0.99-1.20 | 0.087 |
| BMI | 0.98 | 0.96-0.99 | 0.024 | 0.98 | 0.96-0.99 | 0.014 |
| Smoking | 1.13 | 0.97-1.32 | 0.128 | - | - | - |
| FDA | 1.17 | 1.08-1.26 | <0.000 | 1.16 | 1.08-1.25 | <0.000 |
| CRAE | 0.9 | 0.83-0.98 | 0.01 | 0.93 | 0.86-1.00- | 0.062- |
| CRVE | - | - | - | 0.94 | 0.86-1.03 | 0.166 |
| ApoE4 | 1.85 | 1.60 – 2.14 | <0.000 | 1.77 | 1.52-2.05 | <0.000 |
| AD |  |  |  |  |  |  |
| age | 0.92 | 0.89-0.96 | <0.000 | 1.08 | 1.06-1.10 | <0.000 |
| sex | - | - | - | 1.27 | 1.01-1.58 | 0.039 |
| Duration with T2D | 1.01 | 1.00-1.03 | 0.079 | - | - | - |
| Previous CVD | 0.72 | 0.56-0.94 | 0.014 | 0.62 | 0.48-0.80 | <0.000 |
| SBP | 0.98 | 0.97-0.99 | 0.002 | 0.99 | 0.98-0.99 | 0.003 |
| Total Cholesterol | 1.12 | 0.97-1.28 | 0.124 | 1.13 | 0.99-1.28 | 0.06 |
| BMI | - | - | - | 0.98 | 0.96-1.00 | 0.17 |
| Smoking | 1.13 | 0.97-1.32 | 0.128 | 0.86 | 0.69-1.08 | 0.19 |
| FDV | 0.85 | 0.75-0.96 | 0.009 | 0.85 | 0.75-0.96 | 0.009 |
| FDA | 1.33 | 1.16-1.52 | <0.000 | 1.32 | 1.15-1.52 | <0.000 |
| CRVE | 0.87 | 0.78-0.97 | 0.013 | 0.88 | 0.79-0.98 | 0.025 |
| E4 | 2.12 | 1.75-2.57 | <0.000 | 2.1 | 1.69-2.48 | <0.000 |
| VD |  |  |  |  |  |  |
| Age | 0.94 | 0.90-0.98 | 0.009 | 1.11 | 1.08-1.14 | <0.000 |
| Duration with T2D | 1.02 | 1.00-1.03 | 0.093 | - | - | - |
| Previous CVD | 1.24 | 0.94-1.64 | 0.132 | - | - | - |
| GHB | 1.31 | 1.15-1.49 | <0.000 | 1.21 | 1.05-1.40 | 0.009 |
| smoking | 1.39 | 1.06-1.82 | 0.017 | - | - | - |
| FDV | 1.22 | 1.07-1.40 | 0.004 | 1.22 | 1.08-1.38 | 0.002 |
| CRAE | 0.88 | 0.77-1.02 | 0.079 | 0.92 | 0.81-1.04 | 0.185 |
| E4 | 1.44 | 1.10-1.90 | 0.004 | 1.41 | 1.06-1.86 | 0.016 |

GHB Glycated Haemoglobin

For AD we found increasing venular (FDV) and increasing arteriolar (FDA) fractal dimensions associated with a reduced (csHR 0.85; 0.74-0.96) and increased (csHR 1.33; 1.16-1.52) risk respectively. Accounting for competing risks of death did not appreciably change these associations. The ratio of FDA to FDV was also independently significant (1.24; 1.10-1.35). Wider venules indicated by an increased CRVE (0.87; 0.78-0.97) was significantly and independently associated with reduced risk. Again, accounting for competing risk of death did not modify this association.

For VD the associations were markedly different compared to findings for AD. In this case increased FDV alone was associated with increased risk of VD (csHR 1.22, 1.07-1.40). Again this did not change accounting for competing risks of death. There was no indication that the ratio of FDA to FDV was important in this setting. Although CRAE remained in the model indicating a reduced risk it did not achieve significance in either the Cox model or the competing risks model. Measures of tortuosity were dropped from all of the models and therefore did not appear to have an impact on dementia risk in our study.

Table 3 provides the results of our ROC analysis. We found that for each of ACD, AD and VD there was a small increase in the AUC when RVMs were included in the model in addition to genetic information (ApoE4 genotype) and clinical covariates compared to the AUC when only considering genetic and clinical covariates. However, this increase only reached significance in the larger ACD population where the AUC using clinical covariates with genetic data alone was 0.7855 and with inclusion of RVMs it was 0.7896, (p=0.022). Table 4 provides the NRI, its components NRI*e* and NRI*ne* as well as the IDI metrics for the same comparison. Supplementary table 1 provides the full NRI tables. Significant increases in both IDI and NRI were found for all models with inclusion of RVMs. However, it can be seen from the negative NRI*ne* for ACD and VD that this was at the expense of an increase in upgrading non-events to an increased risk category.

**Table 3.** AUC changes with inclusion of RVMs.

|  | Model with RVMs |  | Model without RVMs |  | p |
| --- | --- | --- | --- | --- | --- |
|  | AUC | 95% CI | AUC | 95%CI |  |
| **ACD** | 0.7896 | 0.7731-0.8060 | 0.7855 | 0.7689-0.8022 | **0.022** |
| **VD** | 0.7831 | 0.7566-0.8095 | 0.7769 | 0.7498-0.8041 | 0.090 |
| **AD** | 0.7759 | 0.7532-0.7986 | 0.7685 | 0.7459-0.7910 | 0.094 |

AUC = Area under ROC curve

**Table 4.** Net Reclassification Improvement (NRI) and Integrated Discrimination Improvement (IDI) for inclusion of RVMs.

|  |  | **NR<sub>Ie</sub>** | **NR<sub>Ine</sub>** | **Estimate** | **SE** | **P value** |
| --- | --- | --- | --- | --- | --- | --- |
| **ACD** | **NRI** | 0.05227 | -0.00160 | 0.05067 | 0.02102 | 0.016 |
|  | **IDI** | - | - | 0.00651 | 0.00175 | <0.000 |
| **AD** | **NRI** | 0.04762 | 0.00139 | 0.04901 | 0.02245 | 0.020 |
|  | **IDI** | - | - | 0.00631 | 0.00166 | <0.000 |
| **VD** | **NRI** | 0.052632 | -0.01057 | 0.04206 | 0.01865 | 0.024 |
|  | **IDI** | - | - | 0.00387 | 0.0011 | <0.000 |

## Discussion

To the best of knowledge, we have undertaken the largest prospective study to date of the association of three previously established and widely investigated RVMs, namely, venular and arteriolar calibre, tortuosity and fractal dimension with incident dementia. We found that an increase in retinal arteriolar fractal dimension (FDA) and a reduced arteriolar calibre (CRAE) were associated with increased risk of developing ACD with a median of just less than 10 years in the future. Using our previously validated methodology (10) we have separately considered the two major subtypes of dementia, AD and VD. Here we found marked differences in the direction of association of arteriolar and venular fractal dimension (FDV) between these sub-types. Increases in FDA and FDV were independently associated with respectively, increased and decreased risk of AD. On the other hand, for VD, FDA had no evidence of an association whereas increased FDV was associated with increased risk.

With respect to retinal vascular calibre, we found that whereas increased CRAE was associated with a reduced risk of VD, although this did not reach significance and was further attenuated when accounting for competing risk of death, wider venules (i.e. CRVE) was associated with a reduced risk of AD. These findings were all independent of a comprehensive range of clinical covariates and also independent of ApoE4 genotype which, other than age, is the strongest known predictor of dementia (17). Measures of vascular tortuosity were not selected in any of the models and this appeared to be mainly due to the simultaneous inclusion of other RVMs in our stepwise models indicating a degree of interdependence and co-linearity as indicated by our correlation matrix. Because RVMs have been associated with a range of vascular-based health outcomes (13), indicating the possibility our findings may be influenced by healthy survivor bias, we undertook a competing risk of death which did not appreciably modify these findings. Finally, we have also demonstrated that RVMs used for this study provide small increments in prediction accuracy for dementia beyond ApoE4 genotype and a broad range of relevant clinical covariates. For the larger study of ACD, the increase in AUC was significant whereas for the smaller sub-studies of AD and VD, although the increases were of a similar magnitude, due to the wider confidence intervals the increase did not achieve significance. A previous case control study has also demonstrated that RVMs can increase the AUC for prediction of AD (18). While this study considered a larger number of RVMs, it did not consider their independence from Apo E4. While evaluation of risk prediction models requires measures of model performance, like the ROC metric, its interpretation is based upon the ability of the predicted probabilities to correctly discriminate between randomly selected diseased and non-diseased subjects. It is not uncommon for the AUC to show only small and non-significant improvements, even for a predictor variable that makes a significant independent contribution. We therefore incorporated previously proposed alternative methods for evaluating the utility of a new marker, especially for cohort studies, that included event-specific reclassification tables and integrated discrimination improvement (19). When using these metrics, we found that RVMs significantly improved risk stratification for each of ACD, AD and VD. This further highlights the concept of the retina potentially containing clinically useful information for predicting future brain health and the potential value of developing further disease-specific imaging biomarkers from retinal images. Importantly, however, in the case of ACD and VD, the positive significant NRI was at the expense of an increased number of non-events being upgraded to a higher risk category. As has previously been indicated (15) when accounting for population prevalence of dementia the NRI value in this case becomes negative with potential implications for use of RVMs for selecting patients for future trials for example.

Our findings in relation to retinal vessel calibre and tortuosity seem to be consistent with previous studies where generally reduced retinal vascular calibres have been found in relation to dementia and findings for vessel tortuosity have been inconsistent (3,4). In contrast, some previous studies that have measured retinal fractal dimensions separately for both arterioles and venules have found both reduced FDA and FDV being associated with increased risk of Alzheimer’s dementia (20–23). However, other studies that considered MRI imaging aspects of brain health indicated a less consistent relationship, with opposing associations of FDA and FDV (24,25) similar to our findings. VD is largely defined by imaging evidence of cerebrovascular disease, often manifesting as lacunar stroke. There have also been conflicting associations of retinal vascular fractal dimensions with cerebrovascular disease for example with increased (26), decreased (27) or no association (28) with overall fractal dimensions.

There are several reasons why our results may differ from previous studies. Firstly, the design of many studies has been case control and of relatively small size, with the inherent challenges with respect to bias and confounding. We have used a large prospective cohort study design considering the association of RVMs many years in advance of a dementia diagnosis. The only other large prospective study we are aware of is the Rotterdam study which to our best knowledge has to date only considered measures of retinal vascular calibre (7). Interestingly however, this study, in contrast to ours, found increased venular calibre was associated with increased dementia risk. Secondly, unlike these previous studies, our study has been conducted entirely in patients with type 2 diabetes. Given the well-established global microvascular disease associated with diabetes, the relationship between retinal RVMs, brain health and dementia outcomes may be different in this patient group. A further potential issue is the fact that in our study we have used EMR to define our dementia cases whereas smaller case control studies tend to have used directly ascertained and assessed Alzheimer’s dementia cases. Nevertheless, in our previous study we were able to demonstrate that a gene score specific for Alzheimer’s disease was strongly associated with AD cases adjudicated by our EMR algorithm, whereas we found no association of this score with VD cases also adjudicated by our algorithm (10). Furthermore, in that study, as well as in the present study, Apo E4 genotype was significantly associated with both AD and VD. Together these are indicators that the pathoaetiological basis of our case definitions in this study are robust. Finally, there are many different algorithm-based approaches to measurement of RVMs and particularly for FD (29) and these may have a profound impact on associations observed in different studies.

A recent study has further highlighted the potential of retinal fundus images as a source of imaging biomarkers to predict disease state and risk (30). In this case a deep learning approach achieved predictions for cardiovascular disease similar to conventional clinical risk screening but using the retina alone. While such an approach may increase the accuracy of prediction, it does not yield features that can be clinically interpreted. However, that study underscored the retinal vasculature as being important in discriminating disease states. In our study the RVMs employed are not necessarily specific to dementia, i.e. have neither been identified as such clinically or pathologically or specifically developed for example through a targeted machine learning approach. Hence, our results suggest that further increases in prediction accuracy may be possible if retinal biomarkers specific to dementia could be identified. This would have important implications for future dementia research aimed at developing therapeutic interventions for reducing dementia risk.

Microvascular integrity is emerging as an important aspect of the neurodegenerative process and brain health (31). The fractal dimension of a complex branching pattern, such as the retinal vascular tree, can be considered as a measure of how complex the pattern is and how it fills in the surrounding space (32); it can be therefore regarded as a measure of the extent of tissue vascularisation and, by implication, its vascular health and resilience. Retinal vascular fractal dimensions may be important indicators of susceptibility to neurodegenerative disease (33). In this context it is interesting to speculate how our findings of the relative balance between FDV and FDA predispose to one type of disease compared to another. Intra-brain vascular dysregulation is an early pathological event during AD development (34) and the role of venular networks in the perivascular clearance of Aβ is emerging as potentially important (35). Our finding of an increased ratio FDV to FDA being associated with increased future AD risk together with a significantly reduced venular calibre may indicate an underlying suboptimal venular network with a predisposition to impaired Aβ clearance by the venular network. The extent to which these findings indicate an inherent developmental or genetic predisposition or are influenced by other environmental exposures cannot be determined from this work, although the independence of the findings from clinical and genetic covariates perhaps indicates an endogenous predisposition. As the number of genomic studies of retinal vascular parameters increase (36) this be approached with genomic instruments in future studies.

## Conclusions

In our large prospective study in patients with type 2 diabetes we have found that RVMs predict future incidence of dementia independently of a wide range of clinical and genetic risk factors. This indicates the possibility that inter-individual developmental or acquired differences in microvascular parameters within the retina can be measured and used to predict susceptibility in patients with diabetes. Further development of such an approach may in future be used in developing strategies for dementia prevention.

## Data Availability

The data used in this research is available from the University of Dundee as a component of the GoDARTS and SHARE combined bio resource.

## Acknowledgements

The authors acknowledge all the GoDARTS participants and all personnel at the University of Dundee who have assisted in the development and maintenance of the GoDARTS bioresource. We also acknowledge the Health Informatics Centre at the University of Dundee for the provision of anonymised clinical data from NHS Tayside the original data owner.

---

**Table S1.** NRI Tables for inclusion of RVMs.

| **ACD** |  | **<10%** | **10-20%** | **>=20%** | **Total** |
| --- | --- | --- | --- | --- | --- |
| **1** | **<10%** | 120 | 27 |  | 147 |
|  | **10-20%** | 17 | 142 | 26 | 185 |
|  | **>=20%** |  | 13 | 95 | 108 |
|  | **Total** | **137** | **182** | **121** | **440** |
| **0** | **<10%** | 4,284 | 134 |  | 4,418 |
|  | **10-20%** | 139 | 613 | 76 | 828 |
|  | **>=20%** | 1 | 61 | 304 | 366 |
|  | **Total** | **4,424** | **808** | **380** | **5,612** |
| **AD** |  |  |  |  |  |
| **1** | **<10%** | 175 | 23 |  | 198 |
|  | **10-20%** | 12 | 70 | 9 | 91 |
|  | **>=20%** |  | 5 | 21 | 26 |
|  | **Total** | **187** | **98** | **30** | **315** |
| **0** | **<10%** | 4,879 | 122 |  | 5,001 |
|  | **10-20%** | 136 | 439 | 43 | 618 |
|  | **>=20%** | 1 | 36 | 81 | 118 |
|  | **Total** | **5,016** | **597** | **124** | **5,737** |
| **VD** |  |  |  |  |  |
| **1** | **<10%** | 165 | 12 |  | 177 |
|  | **10-20%** | 2 | 26 | 1 | 29 |
|  | **>=20%** |  |  | 3 | 3 |
|  | **Total** | **167** | **38** | **4** | **209** |
| **0** | **<10%** | 5,478 | 91 |  | 5,569 |
|  | **10-20%** | 45 | 204 | 19 | 268 |
|  | **>=20%** |  | 3 | 23 | 26 |
|  | **Total** | **5,523** | **298** | **42** | **5,863** |

**Figure S1:**
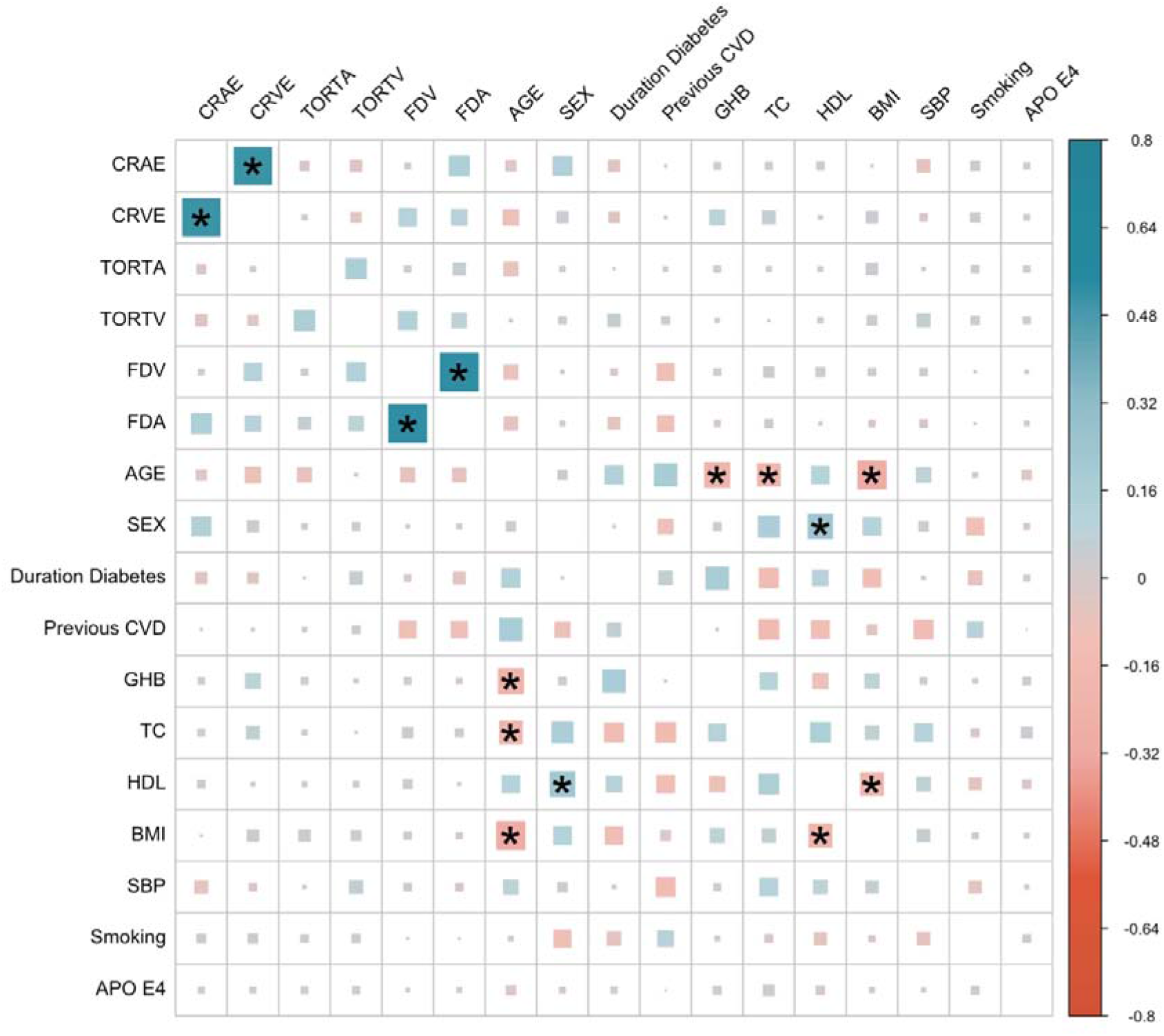
**Correlation matrix for all variables used in the analysis. Direction and magnitude of correlation given by colour scale and magnitude augmented by size. Significant correlations are denoted by \***. **GHB = Glycated Haemoglobin, TC=Total Cholesterol, HDL = HDL Cholesterol, BMI = Body Mass Index, SBP = Systolic Blood Pressure.**

## References

1. London A, Benhar I, Schwartz M. The retina as a window to the brain - From eye research to CNS disorders. Nat Rev Neurol [Internet]. 2013;9(1):44–53. Available from: http://dx.doi.org/10.1038/nrneurol.2012.227

2. Mehta D, Jackson R, Paul G, Shi J, Sabbagh M. Why do trials for Alzheimer’s disease drugs keep failing? Expert Opin Investig Drugs. 2017;26(6):735–9.

3. Wu H, Wang C, Chen C, Xu X, Zhu Y, Sang A, et al. Association between retinal vascular geometric changes and cognitive impairment: A systematic review and meta-analysis. J Clin Neurol. 2020;16(1):19–28.

4. McGrory S, Cameron JR, Pellegrini E, Warren C, Doubal FN, Deary IJ, et al. The application of retinal fundus camera imaging in dementia: A systematic review. Alzheimer’s Dement (Amsterdam, Netherlands) [Internet]. 2017 [cited 2018 Jan 13];6:91–107. Available from: http://www.ncbi.nlm.nih.gov/pubmed/28229127

5. Klaver CC, Ott A, Hofman A, Assink JJ, Breteler MM, de Jong PT. Is Age-related Maculopathy Associated with Alzheimer’s Disease11? Am J Epidemiol. 1999;150(9):963–8.

6. Schrijvers EMC, Buitendijk GHS, Ikram MK, Koudstaal PJ, Hofman A, Vingerling JR, et al. Retinopathy and risk of dementia: The rotterdam study. Neurology. 2012;79(4):365–70.

7. de Jong F, Schrijvers E, Ikram M, Koudstaal P, De Jong P, Hofman A, et al. Retinal vascular caliber and risk of dementia. Neurology. 2011;76(9):816–21.

8. Xue M, Xu W, Ou Y-N, Cao X-P, Tan M-S, Tan L, et al. Diabetes mellitus and risks of cognitive impairment and dementia: A systematic review and meta-analysis of 144 prospective studies. Ageing Res Rev [Internet]. 2019;55(July):100944. Available from: https://doi.org/10.1016/j.arr.2019.100944

9. Kalaria RN. Neuropathological diagnosis of vascular cognitive impairment and vascular dementia with implications for Alzheimer’s disease. Acta Neuropathol [Internet]. 2016 May [cited 2019 Jan 3]; 131(5):659–85. Available from: http://www.ncbi.nlm.nih.gov/pubmed/27062261

10. Doney ASF, Bonney W, Jefferson E, Walesby KE, Bittern R, Trucco E, et al. Investigating the Relationship Between Type 2 Diabetes and Dementia Using Electronic Medical Records in the GoDARTS Bioresource. Diabetes Care [Internet]. 2019 Oct 1 [cited 2019 Oct 7];42(10):1973–80. Available from: http://www.ncbi.nlm.nih.gov/pubmed/31391202

11. Hébert HL, Shepherd B, Milburn K, Veluchamy A, Meng W, Carr F, et al. Cohort Profile: Genetics of Diabetes Audit and Research in Tayside Scotland (GoDARTS). Int J Epidemiol [Internet]. 2017 Sep 7 [cited 2018 Jan 16]; Available from: http://academic.oup.com/ije/article/doi/10.1093/ije/dyx140/4107246/Cohort-Profile-Genetics-of-Diabetes-Audit-and

12. Perez-Rovira A, MacGillivray T, Trucco E, Chin KS, Zutis K, Lupascu C, et al. VAMPIRE: Vessel assessment and measurement platform for images of the REtina. In: 2011 Annual International Conference of the IEEE Engineering in Medicine and Biology Society [Internet]. IEEE; 2011 [cited 2018 Jan 13]. p. 3391–4. Available from: http://ieeexplore.ieee.org/document/6090918/

13. MacGillivray TJ, Trucco E, Cameron JR, Dhillon B, Houston JG, van Beek EJRR. Retinal imaging as a source of biomarkers for diagnosis, characterization and prognosis of chronic illness or long-term conditions. Br J Radiol [Internet]. 2014 Aug [cited 2018 Jan 13];87(1040):20130832. Available from: http://www.ncbi.nlm.nih.gov/pubmed/24936979

14. Pencina MJ, D’Agostino RB, Pencina KM, Janssens ACJW, Greenland P. Interpreting incremental value of markers added to risk prediction models. Am J Epidemiol. 2012;176(6):473–81.

15. Kerr KF, Wang Z, Janes H, Mcclelland RL, Pepe MS. Net Reclassification: critical review. 2015;25(1):114–21.

16. Byberg L, Gedeborg R, Michae K. Useful tests of usefulness of new risk factors⍰: Tools for assessing reclassification and discrimination. 2011;(December 2010):439–41.

17. Michaelson DM. APOE ε4: The most prevalent yet understudied risk factor for Alzheimer’s disease. Alzheimer’s Dement [Internet]. 2014 Nov 1 [cited 2019 Jan 7];10(6):861–8. Available from: https://www.sciencedirect.com/science/article/pii/S1552526014024996?via%3Dihub

18. Cheung CYL, Ong YT, Ikram MK, Ong SY, Li X, Hilal S, et al. Microvascular network alterations in the retina of patients with Alzheimer’s disease. Alzheimer’s Dement [Internet]. 2014;10(2):135–42. Available from: http://dx.doi.org/10.1016/j.jalz.2013.06.009

19. Pencina MJ, Sr RBDA, Jr RBDA, Vasan RS. Evaluating the added predictive ability of a new marker11: From area under the ROC curve to reclassification and beyond. 2008;(June 2007):157–72.

20. Frost S, Kanagasingam Y, Sohrabi H, Vignarajan J, Bourgeat P, Salvado O, et al. Retinal vascular biomarkers for early detection and monitoring of Alzheimer’s disease. Transl Psychiatry [Internet]. 2013 Feb 26 [cited 2019 Jan 17];3(2):e233. Available from: http://www.ncbi.nlm.nih.gov/pubmed/23443359

21. Cheung CY, Ong S, Ikram MK, Ong YT, Chen CP, Venketasubramanian N, et al. Retinal Vascular Fractal Dimension Is Associated with Cognitive Dysfunction. J Stroke Cerebrovasc Dis [Internet]. 2014 Jan 1 [cited 2019 Jan 17];23(1):43–50. Available from: https://www.sciencedirect.com/science/article/pii/S1052305712003096?via%3Dihub

22. Williams MA, McGowan AJ, Cardwell CR, Cheung CY, Craig D, Passmore P, et al. Retinal microvascular network attenuation in Alzheimer’s disease. Alzheimer’s Dement Diagnosis, Assess Dis Monit [Internet]. 2015;1(2):229–35. Available from: http://dx.doi.org/10.1016/j.dadm.2015.04.001

23. Jung NY, Han JC, Ong YT, Cheung CY lui, Chen CP, Wong TY, et al. Retinal microvasculature changes in amyloid-negative subcortical vascular cognitive impairment compared to amyloid-positive Alzheimer’s disease. J Neurol Sci [Internet]. 2019;396(April 2018):94–101. Available from: https://doi.org/10.1016/j.jns.2018.10.025

24. Crystal HA, Holman S, Lui YW, Baird AE, Yu H, Klein R, et al. Association of the fractal dimension of retinal arteries and veins with quantitative brain MRI measures in HIV-infected and uninfected women. PLoS One. 2016;11(5):1–11.

25. Hilal S, Ong YT, Cheung CY, Tan CS, Venketasubramanian N, Niessen WJ, et al. Microvascular network alterations in retina of subjects with cerebral small vessel disease. Neurosci Lett [Internet]. 2014;577:95–100. Available from: http://dx.doi.org/10.1016/j.neulet.2014.06.024

26. Cheung N, Liew G, Lindley RI, Liu EY, Wang JJ, Hand P, et al. Retinal fractals and acute lacunar stroke. Ann Neurol. 2010;68(1):107–11.

27. Doubal FN, MacGillivray TJ, Patton N, Dhillon B, Dennis MS, Wardlaw JM. Fractal analysis of retinal vessels suggests that a distinct vasculopathy causes lacunar stroke. Neurology. 2010;

28. Cheung CYL, Tay WT, Ikram MK, Ong YT, De Silva DA, Chow KY, et al. Retinal microvascular changes and risk of stroke: The Singapore Malay eye study. Stroke. 2013;44(9):2402–8.

29. Huang F, Dashtbozorg B, Zhang J, Bekkers E, Abbasi-Sureshjani S, Berendschot TTJM, et al. Reliability of Using Retinal Vascular Fractal Dimension as a Biomarker in the Diabetic Retinopathy Detection. J Ophthalmol. 2016;2016.

30. Poplin R, Varadarajan A V., Blumer K, Liu Y, McConnell M V., Corrado GS, et al. Prediction of cardiovascular risk factors from retinal fundus photographs via deep learning. Nat Biomed Eng [Internet]. 2018 Feb 19 [cited 2018 Feb 20];1. Available from: http://www.nature.com/articles/s41551-018-0195-0

31. Erdener ŞE, Dalkara T. Small vessels are a big problem in neurodegeneration and neuroprotection. Front Neurol. 2019;10(AUG):1–20.

32. Patton N, Aslam TM, MacGillivray T, Deary IJ, Dhillon B, Eikelboom RH, et al. Retinal image analysis: Concepts, applications and potential. Prog Retin Eye Res. 2006;25(1):99–127.

33. Lemmens S, Devulder A, Van Keer K, Bierkens J, De Boever P, Stalmans I. Systematic Review on Fractal Dimension of the Retinal Vasculature in Neurodegeneration and Stroke: Assessment of a Potential Biomarker. Front Neurosci. 2020;14(January).

34. Iturria-Medina Y, Sotero RC, Toussaint PJ, Mateos-Pérez JM, Evans AC, Weiner MW, et al. Early role of vascular dysregulation on late-onset Alzheimer’s disease based on multifactorial data-driven analysis. Nat Commun. 2016;7(May).

35. Morrone CD, Bishay J, McLaurin J. Potential role of venular amyloid in alzheimer’s disease pathogenesis. Int J Mol Sci. 2020;21(6):1–18.

36. Veluchamy A, Ballerini L, Vitart V, Schraut KE, Kirin M, Campbell H, et al. Novel Genetic Locus Influencing Retinal Venular Tortuosity Is Also Associated With Risk of Coronary Artery Disease. Arterioscler Thromb Vasc Biol. 2019;(December):2542–52.

